# Predicting and Preventing Suicide at Entry to Mental Health Care: A Community-Engaged, Machine Learning Model Implementation

**DOI:** 10.1101/2025.03.30.25324907

**Authors:** Honor Hsin, Santiago Papini, Yun Lu, Heather Clancy, Megan Erion, Carrie Lee, Kimberly Brochard-Des Roches, Jinghui Ju, Kristine Girard, Maria Koshy, Sameer Awsare, Stacy A. Sterling, Vincent X. Liu, Esti Iturralde

**Author notes:** **Description:** A five-step approach to machine learning implementation in suicide prevention illustrates how health systems can bridge individual and population care needs in response to an urgent public health crisis.

## Abstract

Suicide rates in the United States have increased steadily over the past twenty years, a trend coinciding with rising use of mental health services across the country. To help patients before a suicide attempt, health systems must be able to screen for suicide risk and take action at a large scale. Recently, powerful machine learning (ML) models have emerged that can accurately predict suicide attempts by using historical electronic health record (EHR) data, and yet there exists no standardized framework for implementing these models in care delivery. Here we present a case study describing the deployment of a suicide risk prediction model within a large virtual mental healthcare program at Kaiser Permanente Northern California that handles more than 5,000 intake visits per month. Our approach used data science to evaluate model validity for our novel use case (intake visits). We integrated patient and clinician voices to design a model-augmented suicide assessment workflow, which we tested iteratively with continuous input from clinician managers. Understanding the opportunities and pitfalls of model-augmented suicide risk assessment from the perspective of patients and clinicians provided an intuitive framework for mapping clinical actions to potential prediction scenarios. This playbook can be applied to ongoing co-development of ML uses, as part of health systems’ continuous learning initiatives integrating ML to serve today’s public health needs.

**Key Takeaways:**

‒ Machine learning models using electronic health record data are able to predict with high likelihood which patients will attempt suicide in the near future.
‒ To make an impact, these models must be deployed in a way that respects patients’ preferences, avoids reinforcing societal inequities, and mitigates workforce burdens.
‒ We implemented a machine learning model to identify individuals at risk for suicide in a large, virtual mental health program handling more than 5,000 monthly intake visits.
‒ Our data-driven and community-engaged approach validated the use of machine learning to augment suicide risk assessment and catch “silent sufferers,” while guiding the development of a manageable workflow and efficient, responsive clinician training strategies.

## The Challenge

The US suicide rate remains on the rise, with 49,476 lives lost to suicide in 2022.^1^ Interventions at the system level and directly with patients can reduce suicidal behaviors when focused on individuals at elevated risk.^2,3,4^ How to screen effectively for suicide risk is not well understood, however, nor are the potential burdens of this screening.^3^ Machine learning (ML) models have been developed recently from electronic health records (EHR) to predict suicide risk.^5,6,7,8^ Using variables such as past clinical diagnoses, medication history, mental health care utilization, and medical history, these models have demonstrated high accuracy at identifying patients at risk for a suicide attempt within 90 days.^5^ Research has begun to examine the impact of harnessing this prediction approach in care delivery; yet no clear implementation pathways exist given the knowledge gap in burdens of suicide risk screening.^9,10,11,12,13^ There is also concern among implementers that ML models could perpetuate societal inequities if they are trained on data that may be incomplete for populations facing barriers to care.

Kaiser Permanente Northern California (KPNC) serves a diverse population of approximately 4.6 million members, with a prevalence of mental health conditions comparable to US rates.^14^ KPNC has been a pioneer in telemedicine including in telepsychiatry, with video visit availability preceding the COVID-19 pandemic. As far back as 2018, the system provided initial mental health assessments (“intake” appointments) by specialized clinicians working virtually. Leaders recognized value in implementing suicide risk prediction for these virtual intake visits. A mental health intake with a clinician is the first step to establishing an episode of care, which may include therapy, medication management, addiction treatment, and/or intensive care options. In 2020, KP had also implemented a systematic system for measurement-based care. This platform includes a widely-used, brief questionnaire completed by the patient to describe their depression symptoms, the Patient Health Questionnaire-9 or PHQ-9, which includes an item on suicidal thoughts. If significant suicidal ideation is reported, an EHR alert reminds clinicians to complete a suicide risk assessment with the patient during the visit.

Clinical operation leaders determined this would be a high-value opportunity to pilot enhanced suicide prevention efforts. Indeed, the intake visit is a unique point-of-care in a patient’s mental health journey, as over 70% of treatment disengagement occurs after the first or second visit.^15^ Identifying at-risk individuals at intake can help ensure that appropriate suicide risk evaluation and treatment planning take place as patients are connecting to care.

## The Goal

The goal was to develop a feasible, scalable prototype of a clinical workflow incorporating ML-based suicide risk models into suicide prevention efforts at the mental health intake point-of-care.

## The Execution

We aimed to adapt ML modeling to our population’s needs with a five-step implementation approach (Figure 1):

1. Assess whether the predictive model accurately estimates suicide risk for patients seeking new mental healthcare, including for patients in various demographic subgroups;
2. Speak to patients with past experience of suicide-related thoughts to understand what they would need and prefer early in mental health treatment when facing this risk;
3. Gather clinician views on potential benefits and challenges of implementing interventions in response to the risk model to define actions that are impactful and feasible;
4. Assemble a clinical workflow with operational leaders to integrate the above findings within operational constraints and existing clinical guidelines;
5. Educate while iteratively testing the new workflow within the health system, adjusting clinician education and interventions in response to real-time data and feedback.

**Figure 1:**
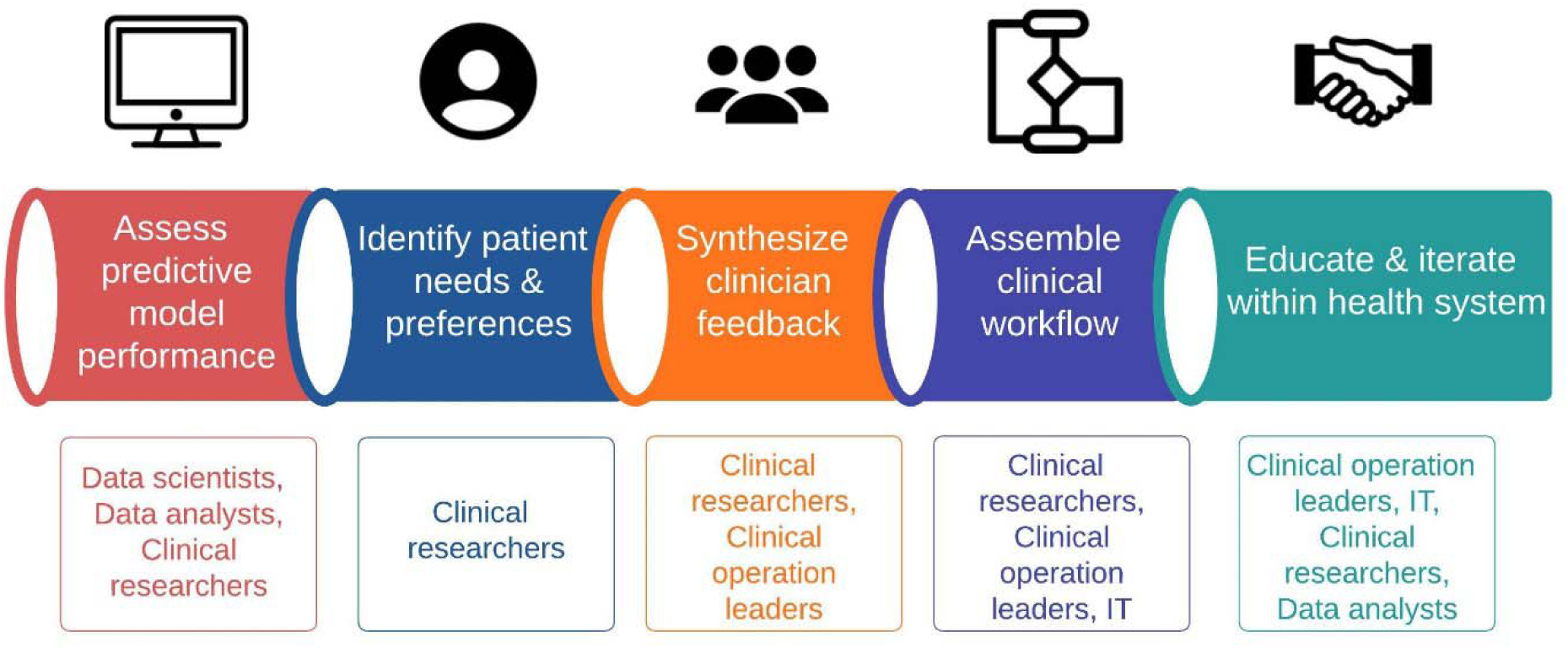
ML implementation pathway for suicide prevention. In each step, actions are summarized in the middle row and critical team members are listed in the bottom row. At KPNC, a predictive analytics core facilitates collaboration among data scientists, clinical researchers, data analysts, and health information technology (IT) specialists. Before the project began, the predictive analytics core was capable of calculating machine learning-based risk scores on individual patient data gated to specific encounter triggers, in near real-time with an approximately 30-minute delay. The team also had the capability to build custom dashboards within the electronic health record (EPIC Systems) for displaying risk score flags customized to pre-set thresholds.

### Step 1: Assess predictive model performance across demographic groups

In 2018, investigators from the Mental Health Research Network published an EHR ML algorithm for predicting risk of suicide attempt within 90 days of a mental health visit.^5^ Our research team evaluated model performance among a narrower population of patients initiating mental health care, and found good performance of an adapted version of this model including across racial and ethnic subgroups.^16^ This was important because algorithmic bias could perpetuate subgroup-specific barriers to mental health care.

### Step 2: Identify patient needs and preferences

Previous studies have highlighted interest among patients in use of ML for suicide risk prediction to direct intervention resources to vulnerable individuals.^17,18,19^ Here, we conducted a design–oriented qualitative study of patient perspectives on suicide risk prediction and interventions, aiming to identify actionable solutions that align with user needs. We interviewed 15 consented KPNC patients identified via EHR who had recently completed a mental health intake visit and self-reported suicide ideation on the PHQ-9 at intake. All participants perceived value in a predictive analytic approach to detect suicide risk. As one young adult participant reflected, a risk model would be beneficial if it helped clinicians to “not miss the silent kids.… You can be going through something, but no one will know.” Notably, participants felt it was important that clinicians’ assessments incorporate additional information beyond what the model could garner from the EHR, such as personal context, past challenges, future goals, and treatment preferences.

We used a “card sort” approach to reveal the underlying values behind participants’ preferences (Figure 2). Interviewees ranked and compared different intervention options, sparking additional exploration of needs.^20^ We asked participants to consider interventions that could be offered during times when they felt at their lowest and to rank these along a scale of helpfulness and a scale of bothersomeness. Compilation of these rankings allowed us to rapidly “fingerprint” general acceptability of interventions, and patients seemed to prefer a clinician contact (via phone or outpatient visit) as the best next step. Underlying patient needs included having choices for treatments, knowledge of their condition, personalization of treatment, and a caring stance of providers. These identified themes of patient need were adopted within potential scripts for clinicians (see Step 4 below).

**Figure 2:**
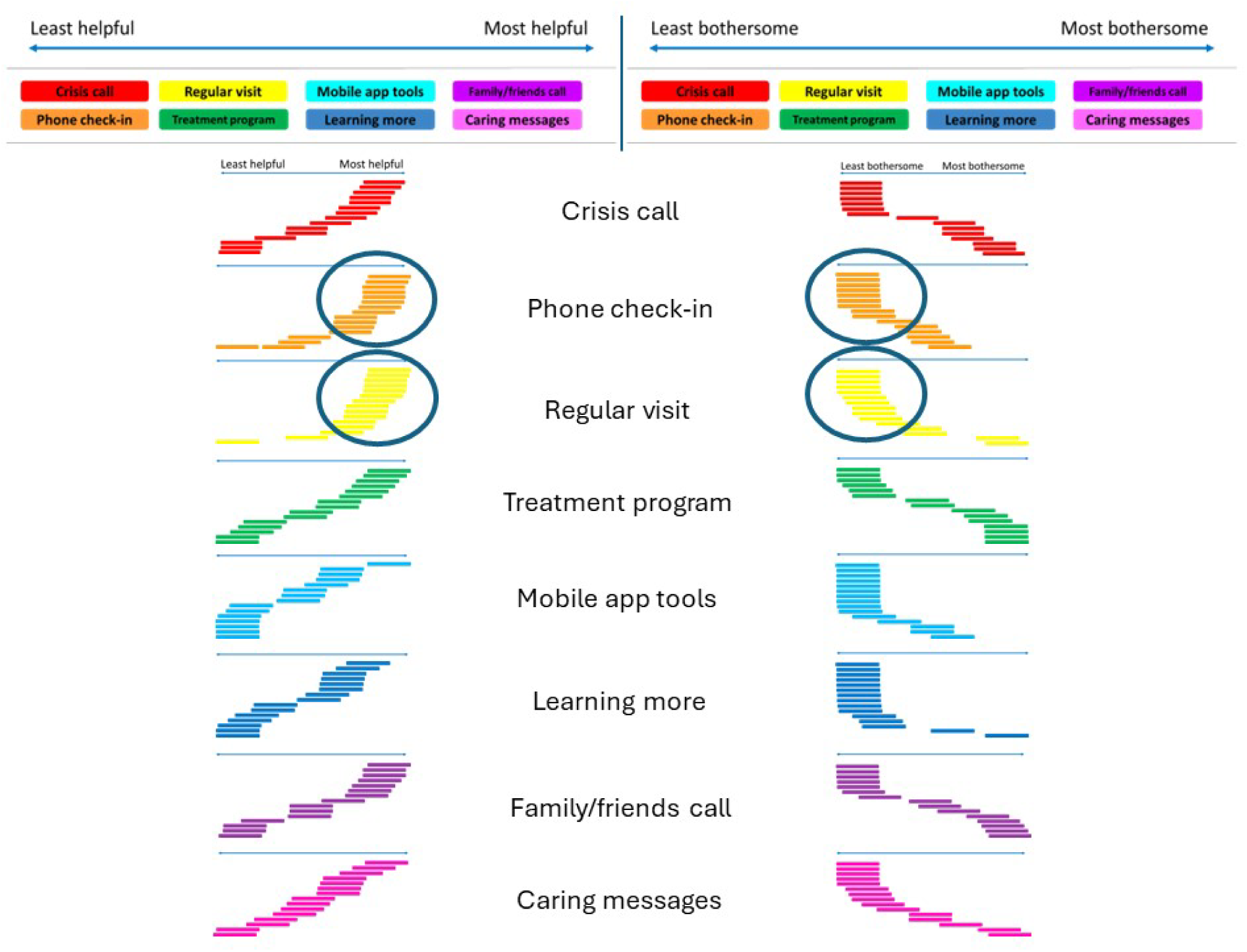
Summary of ML intervention card-sorting exercise with patients. Ages of interviewed patients ranged from 20-81 years, with 6 self-identified males and 9 self-identified females. Self-identified races and ethnicities included White (9), Black (3), Hispanic (2), and multiracial (Black and Native American; 1). Each participant was shown an axis of helpfulness or bothersomeness above, and asked to place one of 8 interventions along these axes. Each placement was then collated together by intervention (one line segment representing a single patient’s placement) to generate a composite preference signature. Interventions included: a call from a crisis hotline (red), a clinician check-in for ∼15 min by phone (orange), a regularly scheduled outpatient clinical visit (yellow), an intensive daily treatment program (green), a referral to mobile app tools (blue), a referral to online resources for more mental health information (indigo), a call from clinicians to family/friends to help the patient (purple), and a message/letter from a clinician conveying caring support (pink). Among participants, a phone check-in or regular visit (such as the intake appointment the participants had just completed) felt the most acceptable to patients in terms of balance between helpfulness and bothersomeness. Other types of interventions appeared to be divided amongst participants, although mobile apps and learning resources seemed least bothersome. These findings structured our workflow approach toward favoring clinician contact via scheduled appointment or phone follow-up in the event of a risk score flag.

### Step 3: Synthesize clinician feedback

Previous studies found general support for suicide risk prediction approaches among providers as well, although some worried about increased time burdens and other clinical demands.^21^ At KPNC, we interviewed 11 consented clinician managers. All clinicians were supportive of suicide risk prediction from EHR data and recognized clinical value. Many volunteered that a risk score flagging approach would fit easily within the current state of suicide alerts that are prompted by computer-based collection of PHQ-9 self-report. Some clinicians felt risk scores offered additional value to the PHQ-9, as they “do not default on just a person’s report in the moment.” Every clinician viewed the next step after seeing a positive ML risk score flag as having the clinician conduct a prioritized suicide risk assessment. They suggested ways to handle potential discrepancies between a ML risk score flag and a self-reported PHQ-9 score. These sample scripts were incorporated into the final workflow (Step 4). Clinicians noted that clinical judgment captured many other factors not readily accessible from the EHR, such as protective factors, current stressors, and “subtle cues” from patient behaviors.

When considering what to do next for model-flagged patients, clinicians favored using their professional experience and judgment to offer patients existing standard-of-care interventions as appropriate. For each potential clinical response, we asked clinicians to describe potential downsides of offering an unnecessary intervention – a hypothetical “false-positive” scenario (Figure 3). A consistent worry was damaging the trust or rapport with the patient. Clinicians also noted the potential for some interventions to be helpful in certain situations, but that for other patients these interventions could be overly intrusive (e.g., a crisis call), or conversely, not supportive enough (e.g., a digital app). Clinicians felt that engaging the patient with the care team through a routine visit was responsive and flexible enough to meet a range of patient needs.

**Figure 3:**
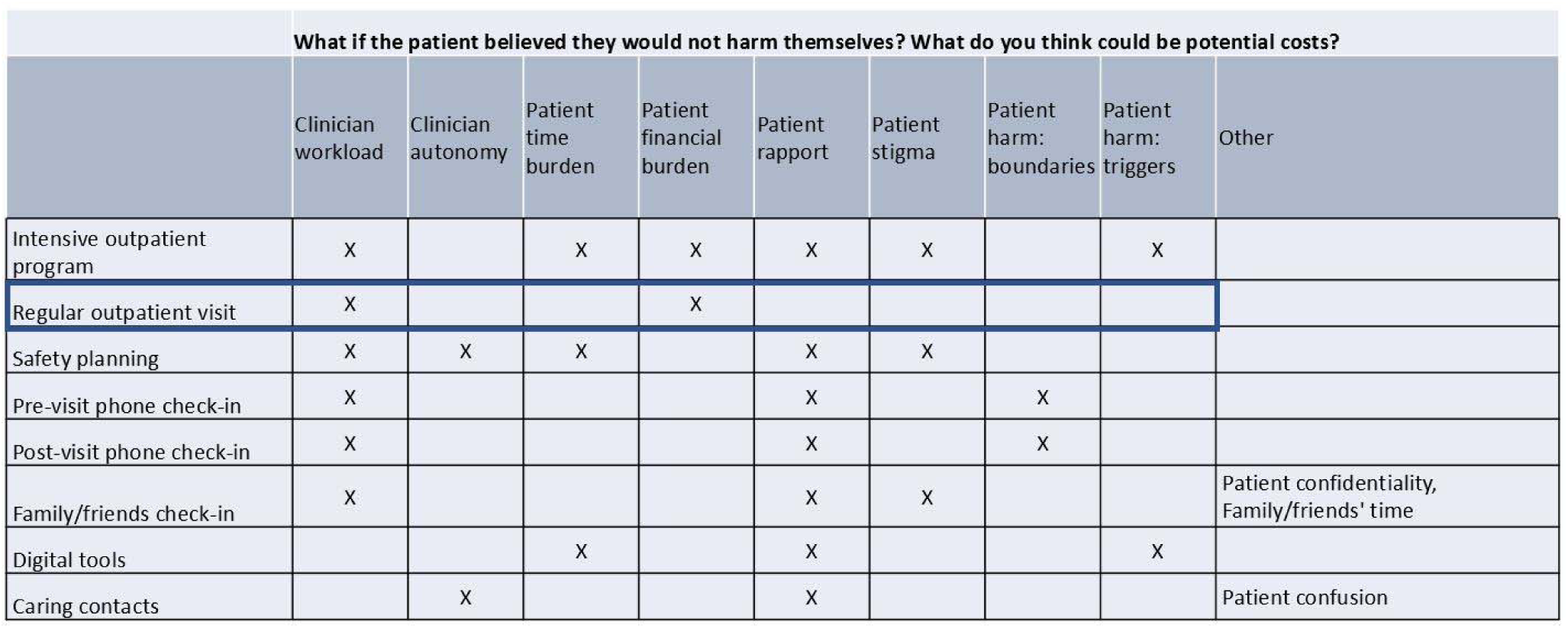
Clinician-identified “cost” burdens (e.g., lost rapport) of ML interventions. Consented clinician managers were interviewed (N = 11). Clinical experience of participants ranged from 5 to 27 years of active practice, and spanned a wide spectrum of clinical settings and levels-of-care, including county clinics, academic centers, federal systems, and private practice. Interventions discussed included (from top row to bottom): intensive outpatient program (multiple visits per week), a regularly scheduled outpatient visit, collaborative completion of a safety plan for patient use, a 15-minute phone check-in before or after an outpatient visit, a brief phone check-in with a patient-identified close contact such as a family member, a referral to digital tools like mobile apps, and a message or letter of caring support sent to the patient. Clinicians identified potential cost burdens of each intervention if offered in a “false-positive” scenario where the ML model flagged a patient who actually did not intend to harm themselves. These burdens were organized thematically as above, and included burdens to clinician workload, clinician autonomy of practice, patient time, patient finances, patient rapport, perceived patient stigma, and patient harms. A regularly scheduled outpatient visit, such as the mental health intake visit, elicited the fewest types of burdens.

### Step 4: Assemble clinical workflow

We next created a workflow based on the above patient and clinician recommendations and incorporated feedback from operational leaders (Figure 4a). The ML risk score flag would augment the self-reported suicide risk alert, with either one triggering the same workflow. The clinician is reminded to conduct a suicide risk assessment at the upcoming visit, and two additional outreach attempts are made in the event of the patient not attending. In the language of electronic circuits,^22^ an ML risk score flag would function “in parallel” to a self-report suicide risk alert, but “in series” to a clinician’s prioritized suicide risk assessment during the intake visit. Such a scenario precisely captured the patient-suggested utility of catching “silent” sufferers.

**Figure 4:**
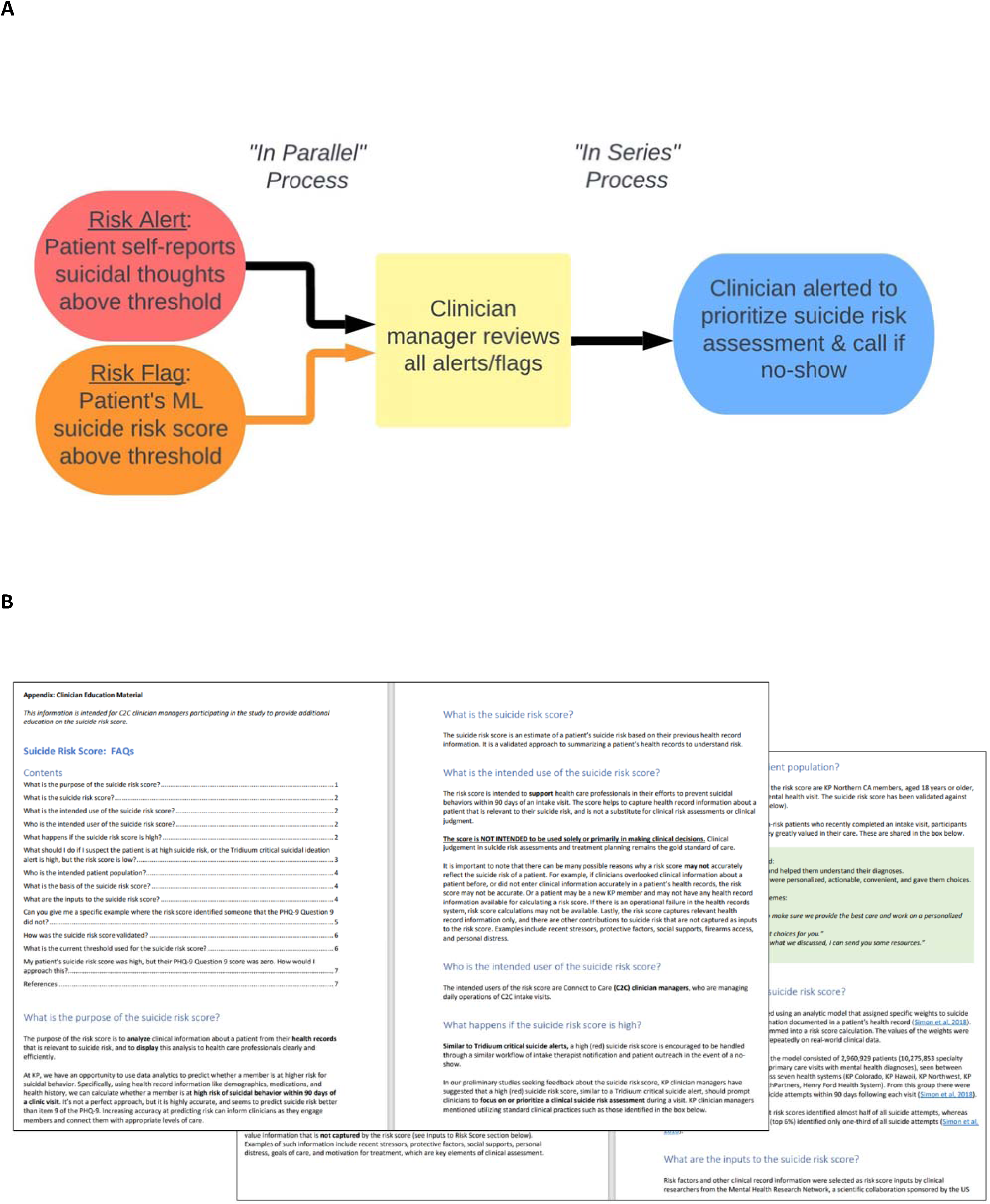
**Clinical workflow and education materials**. Black arrows indicate steps that were already in practice before ML implementation; the orange arrow indicates the new step adopted into the workflow. ML suicide risk score flags for incoming intake visits would be presented to the C2C clinician manager, who would next alert the patient’s intake clinician to conduct a prioritized suicide risk assessment at the scheduled outpatient visit. If a patient no-showed to the visit, the intake clinician would outreach by phone contact for a suicide risk assessment. (b) Sample pages from the clinician education handbook (available upon request). The handbook followed guidelines for user-facing information as outlined in the United States Food and Administration (FDA) September 2022 Guidance for Clinical Decision Support Software.^23^ Additional information requested by clinicians to be included were (a) discussion of pertinent clinical information *not* captured by the suicide risk score, including examples where a patient may be at elevated suicide risk despite a low score, (b) anonymized case examples of two patients with elevated suicide risk scores but denied suicidal thoughts at the visit, to illustrate how a patient’s EHR variables can contribute to their calculated risk.

### Step 5: Educate and iterate within the health system

Our team found it useful to assemble a clinician education handbook that integrated several key components (Figure 4b). In collaboration with clinical leaders who manage intake clinicians, we strategized approaches to clinician education. First, we clarified that clinical judgment remained unchanged, as clinicians would continue to conduct suicide risk assessments according to standard practices. Second, we discussed the risk score from a descriptive rather than a prescriptive approach, which helped to minimize potential concerns of a new class of clinical responsibilities. Third, a systems approach to education was embraced, including a “cascading” ladder from leaders to managers to clinicians to anticipate potential frontline concerns. Our leaders noted that having a strong pre-existing suicide risk management workflow was essential for implementing a new approach that introduced additional risk information.

During implementation, we iterated the ML risk score interface based on user experience (clinician feedback, periodic anonymous surveys) and model metrics (risk score frequencies, direct review of flagged patient cases). Anonymized case descriptions were included in training materials. Daily workload was monitored on a weekly-to-monthly basis.

### The Team

A multidisciplinary team engaged in this effort, including:

- data scientists and analysts, with expertise in healthcare data extraction and predictive analytics to validate the ML model use case,
- clinician scientists, with expertise in qualitative research to extract actionable insights from patient and clinician interviews,
- information technologists to execute and iterate on the creation of an ML-based risk score dashboard,
- clinical operation leaders, from managers to directors, to guide provider education and implementation rollout,
- a strong executive sponsorship group across the areas of mental health, population care delivery, healthcare AI, and health implementation research. We found that all four areas of executive presence were crucial.

### Metrics

From 10/19/23 to 4/1/24, the ML model calculated risk scores of 56,124 intake visits. Mean age of patients was 40.1 ± 15.1 years, with 66.7% identifying as female and 33.3% identifying as male gender. One-quarter of patients identified as Hispanic/Latinx ethnicity, and racial distributions were 41.3% White, 14.4% Asian, 9.4% Black, 4.8% multiracial, 0.6% Native Hawaiian or other Pacific Islander, 0.5% American Indian or Alaskan Native, and 7.4% with missing race information. This demographic breakdown was comparable to the composition of our previous model validation efforts.^16^ Individual case reviews illustrated alignment between clinical judgment and model prediction of risk. The evolution of the total number of flagged cases is illustrated in Figure 5. Lastly, surveys indicated that all consented clinician managers consistently “agreed” or “strongly agreed” that the purpose of the suicide risk score was clear.

**Figure 5:**
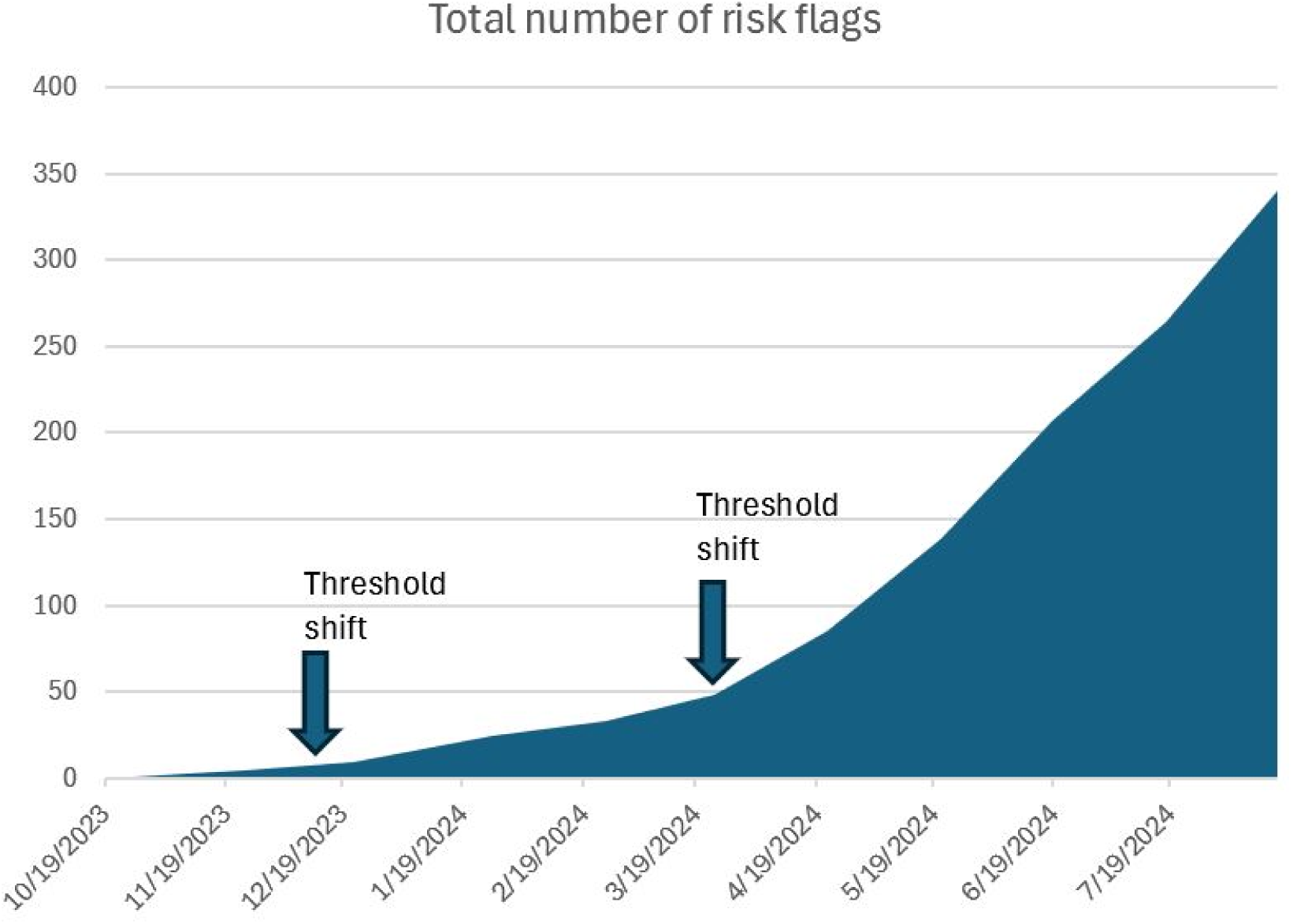
Evolving threshold shift. Total number of patient encounters flagged by the suicide risk score is trended over time. The arrows indicate each time a concerted discussion was made with the clinical operational team to lower the risk score flag threshold.

### Hurdles

Introducing a new population management tool necessitated a balance between outreach and workload. We initially selected an ML risk threshold very conservatively, which appeared to encompass the top-scoring approximately 0.05% of clinic visits. Interestingly, we found that this resulted in so few flags that providers did not perceive a clinical value. We found that operational decisions to set the risk score threshold were best determined by balancing workload estimates with individual review of cases that would become newly flagged. This approach helped our operational leaders to weigh potential costs and benefits to a threshold shift. Two such shifts were approved by operational leaders, with ML risk flags now encompassing the top-scoring approximately 1.5% of clinic visits.

### Where to Start

We describe here an implementation playbook for ML in suicide prevention for a community population. ML in healthcare delivery remains challenging, as traditional pathways to evidence-based practice take time to develop. In the case of suicide prevention, ML can play an augmenting role in the presence of system-level efforts to integrate perspectives of patients and clinicians. Ongoing co-evolution with providers and organizational leaders can further support ML’s contributions toward addressing urgent public health problems.

## Data Availability

All data produced in the present study are within the Kaiser Permanente Northern California (KPNC) healthcare system and are available upon reasonable request to appropriately qualified individuals through the KPNC Division of Research.

## Acknowledgements

We gratefully acknowledge members of the KP Predictive Analytics Core (Cristina Perkins, Michelle Donnelly, Marc Flagg), the Connect-2-Care virtual intake clinicians and clinician managers, and KP Division of Research staff (Melanie Jackson-Morris, Mei-Tsung Lee, Jonathan Lontok, Wei Tao). We also thank Agatha Hinman for copy editing services. This study was approved by the Kaiser Permanente Northern California Institutional Review Board and funded by a Delivery Science Grant from The Permanente Medical Group.

## Notes

### Competing Interest Statement

The authors have declared no competing interest.

### Funding Statement

This study was funded by a Delivery Science Grant from The Permanente Medical Group.

### Author Declarations

The Kaiser Permanente Northern California Institutional Review Board gave ethical approval for this work.

